# Neandertal introgression dissects the genetic landscape of neuropsychiatric disorders and associated behavioral phenotypes

**DOI:** 10.1101/2021.10.08.21264688

**Authors:** Michael Dannemann, Yuri Milaneschi, Danat Yermakovich, Victoria Stiglbauer, Manuel A. Friese, Christian Otte, Brenda W.J.H. Penninx, Janet Kelso, Stefan M. Gold

## Abstract

**Background:** Advances have recently been made in identifying the genetic basis of psychiatric and neurological disorders, however, fundamental questions about their evolutionary origins remain elusive. Here, introgressed variants from archaic humans such as Neandertals can serve as an intriguing research paradigm.

**Methods:** We compared the number of associations for Neandertal variants to the number of associations of frequency-matched non-archaic variants with regard to human CNS disorders (neurological and psychiatric), nervous system drug prescriptions as a proxy for disease, and related non-disease phenotypes in the UK biobank (UKBB), the NESDA cohort and the Biobank Japan.

**Results:** While no enrichment for Neandertal genetic variants were observed in the UKBB for psychiatric or neurological disease categories, we found significant associations with certain behavioral phenotypes including pain, chronotype/sleep, smoking and alcohol consumptions. Several of these associations were also observed in NESDA and the Biobank Japan, suggesting their evolutionary relevance across different ancestry backgrounds. Intriguingly, in some instances, the enrichment signal was driven by Neandertal variants that represented the strongest association genome-wide.

**Conclusions:** Our data suggest that evolutionary processes in recent human evolution like admixture with Neandertals significantly contribute to behavioral phenotypes but not psychiatric and neurological diseases. These findings help to link genetic variants in a population to putative past beneficial effects, which likely only indirectly contribute pathology in modern humans, possibly due to changes in lifestyle and maladaptation.

## Introduction

It has long been known that psychiatric disorders run in families, indicating substantial heredity, but their genetic basis has remained elusive for decades(1). This has changed only fairly recently with the advent of large consortia that have discovered and successfully replicated genome-wide associations of common gene variants (single nucleotide polymorphisms, SNPs) for several disorders including schizophrenia(2) and major depression(3–6).

While evolutionary origins have been postulated for psychiatric(7) and neurological(8) diseases, this hypothesis has not been tested directly. One approach to address this question is to uncover the evolutionary history of phenotype-associated variation. One process through which this variation could have entered a population is through admixture events at some point in the past. Here, introgressed variants from archaic humans can serve as an intriguing research paradigm. After modern humans left Africa more than 60.000 years ago, genetic evidence suggests multiple admixture events ∼55,000 years(9) between modern humans and now extinct archaic humans including Neandertals(10–12) and Denisovans(13,14). After admixture, early events of negative selection removed parts of that archaic DNA such that ∼2% of Neandertal ancestry is still found in all present-day non-Africans(15–17). Some of the remaining Neandertal variants have reached high frequencies in some present-day populations, suggesting that they might have conferred advantages at some point since admixture(12,18,19). Introgressed Neandertal variants are also particularly interesting because they are detectable in all non-African populations(14,20) and their phenotypic correlates can thus be studied across different ancestries (e.g. European, Asian), see **Figure 1A**.

**Figure 1:**
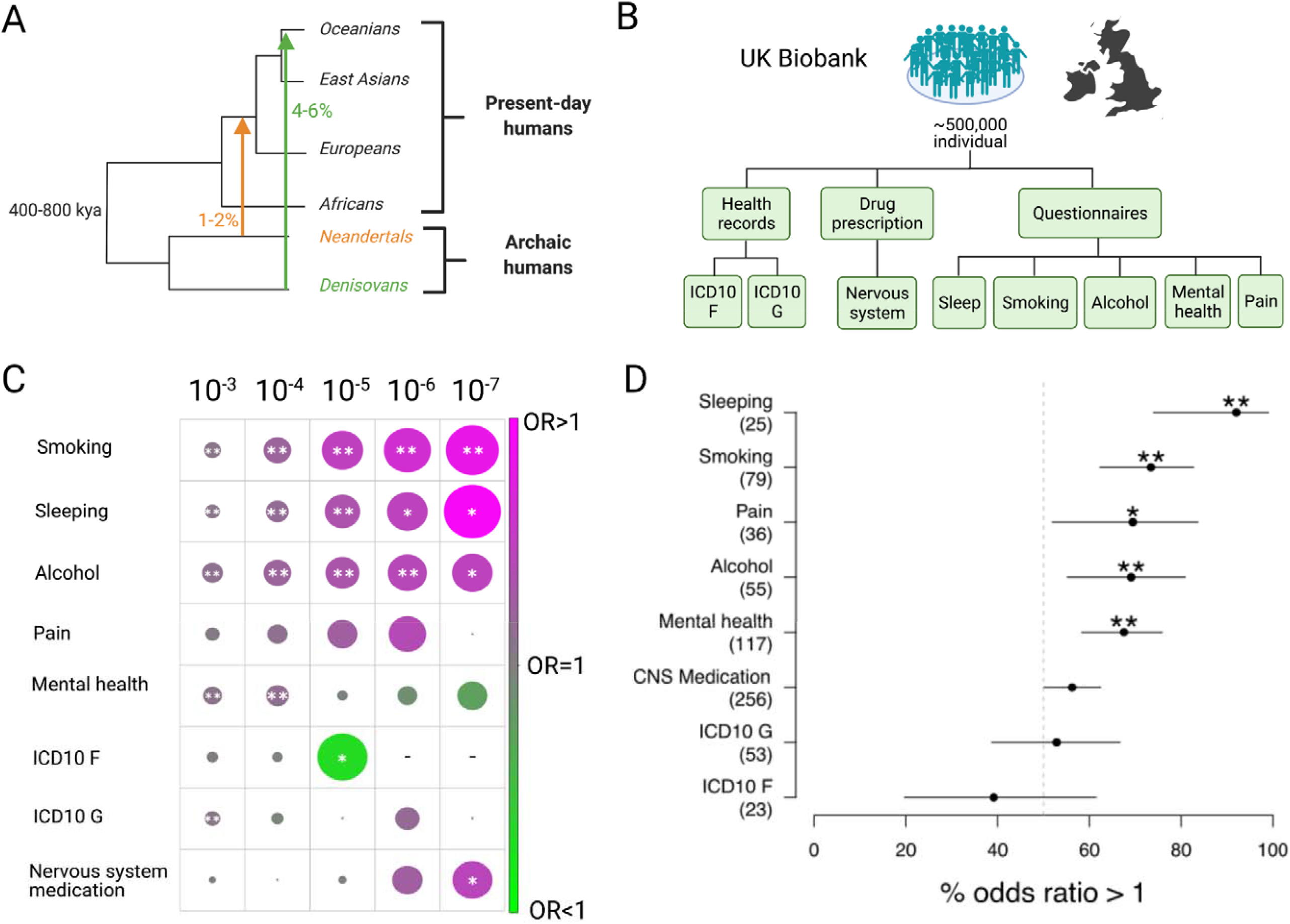
The proportional association of Neandertal DNA with mental and behavioral disease and non-disease phenotypes. **(A)** Some of the genetic variation in present-day people is a result of the admixture with archaic humans such as Neandertals and Denisovans ∼55,000 years ago. Consequently, ∼2% of the genomes of present-day non-Africans are of Neandertal ancestry and an additional 5% of the genomes of Oceanians derived from Denisovans. Approximately 40% of the genome of Neandertals can be reconstructed in people today and tested for its phenotypic potential in genome-wide association studies. **(B)** Overview of UK Biobank GWAS summary statistics for behavioral phenotypes that were included in this study. Included phenotypic information was derived from medical records that include diagnosis and drug prescription information and questionnaires related to behavioral patterns. **(C)** The combined numbers of associations of tag aSNPs with all phenotypes from one of eight tested groups in the UK Biobank (y axis, left) was put in relation to the average of the combined numbers of associations for 1,000 sets of frequency-matched non-archaic tag SNPs. The resulting average odds ratios based on five significance cutoffs (shown on top) are displayed and color-coded and shown in size proportional to the corresponding log of the average odds ratio. Instances where tag aSNPs showed significant deviations from the number of phenotype associations of their non-archaic counterparts were highlighted (*P<0.05, **FDR<0.05). **(D)** The average odds ratio between the number of associations of tag aSNPs compared to the number of associations for 1,000 sets of frequency-matched non-archaic tag SNPs for each individual phenotype from eight phenotype groups from the UK Biobank was calculated. The proportion of odds ratios larger than one for a given group (y axis, number of tests across all significance cutoffs for each group shown in parenthesis), together with the binomial 95% confidence intervals are displayed. Phenotype groups for which this proportion is significantly different from the random expectation of 50% are highlighted (*P<0.05, **FDR<0.05).

In order to identify complex traits that have been significantly influenced by Neandertal DNA, previous studies have compared their numbers of associations of introgressed archaic variants in GWAS to the associations of frequency-matched non-archaic variants. In one study that analyzed health record data in ∼28,000 individuals, the authors demonstrated that neurological and psychiatric disorders showed the highest proportional Neandertal DNA contribution(21). In addition, a second study has shown that among 136 diverse non-disease phenotypes, tested in ∼122,000 individuals of the UK Biobank pilot release, Neandertal DNA was over-proportionally often associated with two mood-related traits, sleeping patterns and smoking status(22). It has been postulated that due to the clinical heterogeneity of complex human disorders, and psychiatric syndromes in particular, careful behavioral phenotyping might yield more robust biological substrates(23). However, a direct comparison of how archaic DNA contributes to diagnostic entities vs. related but non-disease phenotypes is lacking.

To address this question, we conducted a series of analyses of GWAS summary statistics analyses in the UK Biobank (UKBB)(24), comparing associations of Neandertal variants with human CNS disorders, drug prescriptions as a proxy for disease, and non-disease phenotypes (see **Figure 1B**). We also leveraged data from two additional cohorts with sufficient genetic coverage of Neandertal variants and available deep phenotyping, the Netherlands Study of Depression and Anxiety (NESDA)(25) and the Biobank Japan(26). Finally, we identified several Neandertal risk factors that strongly influence these traits and demonstrate that some of them are population-specific.

## Methods

### GWAS cohorts

#### UK Biobank

Summary statistics for 261 genome-wide association studies (GWAS, Table S1) from the UK Biobank(24) were obtained from the Neale lab [http://www.nealelab.is/uk-biobank/]. A detailed description of the analyses underlying these GWAS statistics can be found at http://www.nealelab.is/blog/2017/9/11/details-and-considerations-of-the-uk-biobank-gwas and http://www.nealelab.is/blog/2019/10/24/updating-snp-heritability-results-from-4236-phenotypes-in-uk-biobank.

GWAS have been conducted using 361,194 biobank individuals genotyped at ∼10.8 million SNPs using custom arrays and imputation based on the Haplotype Reference Consortium, the 1000 Genomes Project, and UK10K, and that have passed quality control filters (see Supplementary Methods for a more detailed description).

#### Biobank Japan

We used publicly available summary statistics for the four smoking GWAS from the Biobank Japan(27), that matched phenotypes we analyzed in the UK Biobank The smoking phenotypes have been defined as (1) ever versus never smokers, (2) smoking cessation, (3) age of smoking initiation and (4) quantity of smoking. These GWAS have been conducted in ∼200,000 individuals from the Biobank Japan cohort, with up to 165,436 individuals per GWAS. Individuals were genotyped using custom arrays and imputation based on the 1,000 Genomes, and included 5,826,586 SNPs after application of quality filters (see Supplementary Methods for a more detailed description).

#### The Netherlands Study of Depression and Anxiety (NESDA)

We generated GWAS summary statistics for eight behavioral phenotypes (Table S1). These analyses were conducted in subsets of individuals of the NESDA cohort, ranging between 1,842 and 2,261 individuals. Genotype data was generated based on custom genotyping arrays and subsequent imputation and included 8,657,974 SNPs after quality filtering. Genome-wide association analyses assuming an additive model were carried out using SNPTEST(28). More detailed information on the GWAS can be found in the Supplementary Methods section.

### Definition of Neandertal marker variants

We used previously inferred putative marker SNPs that tag tracts of Neandertal ancestry in the 1,000 Genomes (phase 3)(29,30). Putative introgressed Neandertal variants, referred to as aSNPs, were defined as being (i) absent in the 1,000 Genomes Yoruba population, the population demonstrated to show the lowest levels of Neandertal DNA in this cohort(29) (ii) present in homozygous state in either the Altai or Vindija Neandertal, two high coverage Neandertal genomes(10,11) and (iii) present in at least one 1,000 Genomes non-African individual. Aside from admixture, another genomic feature that could lead to a similar allele-sharing pattern is incomplete lineage sorting (ILS). However, because Neandertal admixture into modern humans is much more recent than the shared common ancestor of Neandertals and modern humans, shared variants that result from introgression are on haplotypes that are much longer than those on which variants resulting from ILS are found. We therefore required (iv) aSNPs to be on haplotypes that exceed the length expected under ILS.

### Testing for the proportional association strength to phenotypes

In order to quantify the association strength of archaic variants we compared their number of significantly associated tag aSNPs to the number of frequency-matched non-archaic tag SNPs following similar approaches that have previously been used for such comparisons(21,22,30,31). These approaches implement measures to account for multiple differences between archaic and non-archaic variants that would otherwise potentially bias this analysis (see Supplementary Methods for a more detailed description).

First, due to the recent admixture ∼55,000 years ago and the separation between modern humans and Neandertals before, introgressed Neandertal DNA is found on haplotypes of tens or even more than 100 thousand base pairs with several aSNPs in high LD. In order to account for linkage disequilibrium (LD) and the differences in the levels of LD between archaic and non-archaic variants we generated sets of SNPs in LD of r^2^>0.5 and selected a random tag SNP. If the set contained aSNPs we annotated it as archaic sets and selected a random tag aSNP to represent the set. Sets without aSNPs were annotated as non-archaic and represented by a random tag SNP. Finally, SNPs without any other variants in LD were defined to be their own tag SNP. In the three cohorts we found 14,839 (UK Biobank), 12,111 (Biobank Japan) and 14,596 (NESDA) tag aSNPs.

We then calculated the number of significant tag aSNP associations based on varying P value cutoffs of 10^−3^, 10^−4^, 10^−5^, 10^−6^, 10^−7^ for the analyses in the Biobank Japan and UK Biobank cohorts and 10^−2^, 10^−3^ for the NESDA cohort. We chose these different cutoffs to account for trait-specific features in individual association analyses, such as heritability or prevalence of the tested trait in the cohort.

A second feature that is specific for aSNPs is its frequency distribution that is linked to the rather low prevalence of Neandertal DNA of ∼2% compared to non-archaic variation in present-day non-African populations. In order to account for frequency-dependent differences between archaic and non-archaic variants and the subsequent differences in detection power we tested for a disproportionate number of tag aSNPs associations by comparing the number tag aSNP association to 1,000 sets of frequency-matched non-archaic tag SNPs.

We then report the average of the 1,000 ratios between the number of tag aSNP associations to the number of association in the random sets in the form of an odds ratio (OR) and empirical P values based on the location of the number of tag aSNP associations within the distribution of associations for the 1,000 random sets. We applied this test to both individual phenotypes and groups of phenotypes. For the latter case we calculated the sum of the numbers of tag aSNP associations across a group of phenotypes and compared it to the sum of numbers of associations of 1,000 random sets of frequency-matched non-archaic tag SNPs.

In order to account for multiple testing we generate false-discovery rates (FDR) using the P value correction approach by Benjamini-Hochberg.

### Functional annotation of Neandertal variants

We defined loci to be Neandertal DNA risk loci if they contained aSNPs with an phenotype association P value below 5×10^−8^ and if these aSNPs were themselves the top association in a given genomic region or in linkage disequilibrium of r2>0.5 with the top associated SNP (Table S2). Frequencies for each candidate aSNP were calculated in 1,000 Genomes populations (phase 3)(29). For each of these aSNP associations we extracted archaic haplotypes with a range that was specified by location of other aSNPs with r2>0.5 with the candidate aSNP (Table S2). We explored GTEx (version 8)(32) for significant eQTLs that were linked to the candidate aSNP or other aSNPs associated with its archaic haplotype. Similarly, we tested whether any candidate aSNP or aSNPs associated with the candidate aSNPs’ haplotype were predicted to modify the amino acid sequence using the ENSEMBLs’ Variant Effect Predictor(33) by extracting all of the aSNPs that were annotated as ‘missense variant’.

## Results

### Diagnostic categories of CNS disorders do not show robust links to Neandertal DNA variants

In the UKBB, our analysis included GWAS summary statistics for 11 mental, behavioral and neurodevelopmental disorders (ICD10 codes F01-F99) and 21 diseases of the nervous system (ICD10 codes G00-G99). The GWAS data was generated based on 361,194 individuals and ∼8 .6 million SNPs with minor allele frequency (MAF) larger than 1% after QC filters on samples and genotypes. (see **Methods** for details).

We annotated putative introgressed Neandertal variants based on previously described marker SNPs, referred to as aSNPs(30). We found and analyzed 197,250 such aSNP in the UK Biobank cohort. We then tested for a disproportionate number of aSNP associations for a given phenotype by calculating the number of LD-corrected tag aSNP associations and comparing it to the numbers of associations in 1,000 random sets of frequency-matched and LD-corrected non-archaic tag SNPs (Methods).

We first applied this method to the two combined groups of disorders. Overall, consistent with the observation by Simonti et al.(21), we found an enrichment for introgressed Neandertal alleles associated with diseases of the nervous system (see **Figure 1C**). However, this was only significant after FDR correction for the least conservative significance cut-off (OR>1, P<0.05 and FDR<0.05 for P value cutoff 10^−3^). Averaged across all significance cutoffs, neither the group of mental, behavioural or neurodevelopmental disorders, nor the group of nervous system diseases codes showed significant enrichment of tag aSNP associations compared to frequency-matched non archaic tag SNPs (see **Figure 1D**). When considering disorders individually, only a few signals appeared, mostly for neuropathies (see **Supplementary Materials**).

As a complementary approach, we next explored medication prescriptions as a proxy for disease in the UKBB. Based on the classification of the World Health Organization, we annotated 626 medications with available data in the UK Biobank (Table S4). When testing whether the cumulative sum of tag aSNP associations across the 96 medications for CNS disorders (category N) differed between tag aSNPs and non-archaic tag SNPs, tag aSNPs showed over-proportional association numbers for one P value cutoff (**Figure 1C**, OR>1, P<0.05 for P value cutoff 10^−7^, Table S1). When averaged across all significance cutoffs, CNS medication did not show significant associations with Neandertal DNA (see **Figure 1D**). However, when breaking down CNS medications into subcategories, we did detect a significant signal for a link between Neandertal DNA and two classes of pain medications (see **Supplementary Materials**).

### Behavioral phenotypes with relevance for mental health show robust enrichment for Neandertal variants

When we explored Neandertal associations with related but “non-disease” behavioral phenotypes, signals became substantially stronger. Questionnaire data were available for numerous phenotypes related to mental health (47 questions), sleep (6), pain (17), smoking (33) and alcohol use (26). We again first quantified the cumulative numbers of tag aSNP associations across GWAS within each of the five groups. We found that GWAS of smoking (all P<0.001 and FDR<0.001, all OR>1, Table S1), sleep traits (all P<0.05 and 4 of 5 FDR<0.05, all OR>1) and alcohol intake (all P<0.05 and 2 of 5 FDR<0.05, all OR>1) showed consistently larger numbers of associations with tag aSNPs when compared to their non-archaic counterparts (Table S1, **Figure 1C**), which were significant across all significance cut-offs. Consequently, these groups of traits showed significant enrichment when averaged across all significance cutoffs (**Figure 1D**).

When analyzing individual GWAS within each of these three groups, we found that six *<u>smoking-related phenotypes</u>* showed notable enrichments; including two describing the number of daily smoked cigarettes (all OR>1, 9 of 10 P<0.05 and 6 of 10 FDR<0.05, Table S1) and another six defining smoking status (8 × P<0.05 and 6 x FDR<0.05, Table S1).

An additional six *<u>alcohol GWAS</u>* showed enrichments (all OR>1 and P<0.05 with one also FDR<0.05). Four of these phenotypes characterized regular intake frequencies for various alcoholic beverages (Table S1), one defining the alcohol drinker status and another one specifying the habit of consuming alcohol during meals. Moreover, four significant phenotypes relate to *<u>chronotype</u>* and one to *<u>sleep duration</u>* (Table S1).

The combined group of *<u>mental health GWAS</u>* showed association enrichment results similar to alcohol, smoking and sleeping habits for the two most relaxed P value cutoffs (OR>1, P<0.01 and FDR<0.05 for 10^−3^ and 10^−4^). With 47 underlying GWAS, the group of ‘mental health’ combines the second largest number of individual GWAS within our tested groups. Among those, 14 GWAS showed at least one enrichment test with OR>1 and P<0.05 and were linked to various mood-related questions (Table S1). The most notable enrichment with odds ratios above 1, three instances of P<0.05 and the only two cases of FDR<0.05 was linked to the length of a depressive episode. The group of mental health phenotype associations also included one with the ‘Longest period of unenthusiasm / disinterest’ where a substantially lower number of associations with tag aSNPs was observed (OR<1 and P<0.05, Table S1).

Finally, while the combined group of *<u>pain phenotypes</u>* showed an average OR>=1 for all tested P value cutoffs, none of these instances reached a significant level of differences between tag aSNP and non-archaic tag SNP associations (Figure 1C-D, Tables S1,S3). On the individual GWAS level, only three tests for pain, one related to each general pain, back pain and knee pain, showed a larger number of tag aSNP associations with P<0.05. One additional GWAS related to long term facial pain even showed lower numbers of associations with Neandertal variants (OR<1 and P<0.05, Table S1).

Taken together, these results indicate a robust enrichment of Neandertal variants associated with 5 groups of behavioral phenotypes in the UKBB: Alcohol consumption, smoking, mental health (specifically mood), chronotype / sleep, and pain.

### Analysis of behavioral phenotype associations in independent cohorts

Next, we explored similar phenotype associations in independent cohorts of diverse ancestry. For this purpose, the Netherlands Study of Depression and Anxiety (NESDA) and the Japanese Biobank provided adequate genetic coverage to analyze tag aSNPs and sufficient depth in the behavioral data.

In NESDA, we were able to probe eight behavioral phenotypes, one for smoking, one for alcohol consumption, two sleep related phenotypes and four mental health phenotypes (Table S1). We applied the same enrichment analysis as in the UKBB, but - due to the reduced association power because of the substantially lower sample sizes in NESDA (N=1,842-2,261) - we adjusted our P value cutoffs to 10^−2^, 10^−3^ (see **Methods**). Out of the 8 tested phenotypes, we found two instances with substantially larger numbers of tag SNP associations: Alcohol intake (OR>1, P<0.05, P value cutoff 10^−2^) and chronotype (OR>1, P<0.05, P value cutoff 10^−3^, Figure 2, Table S1).

**Figure 2:**
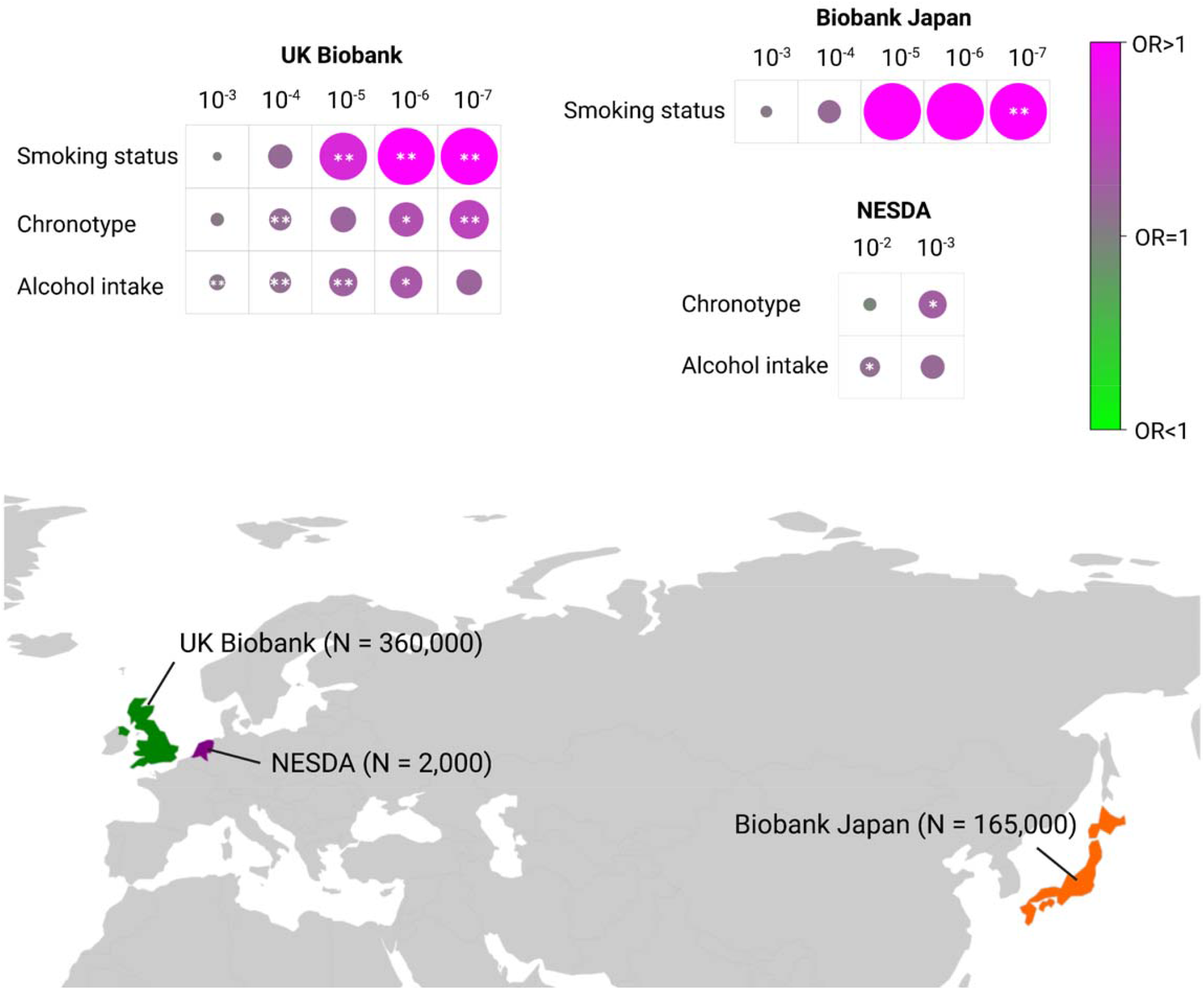
Analysis of Neandertal DNA associations in the Biobank Japan and Netherlands Study of Depression and Anxiety (NESDA). The three phenotype groups with the largest over-proportional Neandertal DNA associations in the UK Biobank cohort were smoking, sleep and alcohol. The individual phenotypes in each group that showed the largest contributions to the respective groups’ enrichment results were smoking status, chronotype and alcohol intake (top left). For these three phenotypes the odd ratio, reflecting the ratio between the number of tag aSNP associations and the average number of associations in 1,000 sets of frequency-matched non-archaic tag SNPs is displayed (color and size indicate the average log odds ratio and direction). Using the same method the three phenotypes were tested for the proportional number of Neandertal DNA associations in the Biobank Japan (for smoking status, top right) and NESDA (for chronotype and alcohol intake, right, middle panel) cohorts. A map illustrating the host countries of each cohort and the sample sizes used for this analysis are displayed in the lower part of the figure.

Both the UK Biobank and NESDA cohorts, include individuals of predominantly European ancestry. However, risk loci for complex traits have often been associated with population-specific genetic variants, a phenomenon that has also been observed for Neandertal DNA(30,34). In order to explore to which extent our results can be translated to cohorts of non-European ancestry, we explored summary statistics from the Biobank Japan. The Biobank provides GWAS summary statistics for four smoking traits derived from ∼165,000 individuals with information about smoking status, daily cigarette consumption and age of onset(27). Again, applying our enrichment method for tag aSNP (Methods) we found that a GWAS for smoking status showed a substantially larger number of Neandertal variants (FDR<0.05, P value cutoff 10^−7^).

Taken together, despite the substantially lower power in NESDA, and the different ancestry in the Japan Biobank, we observed Neandertal enrichment for behavioral phenotypes of smoking, alcohol consumption, and chronotype.

### Exploring the frequency and biological relevance of individual Neandertal variants

In addition, we noted that Neandertal variants were not only significantly enriched among associations for multiple behavioral phenotypes but also contributed high effect risk variants. We found 27 instances linked to 18 independent Neandertal loci that showed genome-wide significant association (P<5×10^−8^), with aSNPs showing the top association or being in high linkage disequilibrium with the lead SNP in a given region (Table S2).

18 of the 27 associations were linked to smoking and sleeping patterns, suggesting that particularly for these phenotypes, Neandertal DNA contributes large effect variants. We also noted that two risk-increasing GWAS lead variants for smoking status in both the UK Biobank (lead aSNP: chr9:136463019_C/A, P=2.7×10^−23^, beta=0.02) and Biobank Japan (lead aSNP: chr8:13289111_C/G, P=6.7×10^−8^, beta=-0.02) were linked to aSNPs (Figure 3A/B, Table S2). Both lead aSNPs showed population-specific frequency differences, a pattern that was highly prevalent among other high-risk Neandertal variants as well (Table S2).

**Figure 3:**
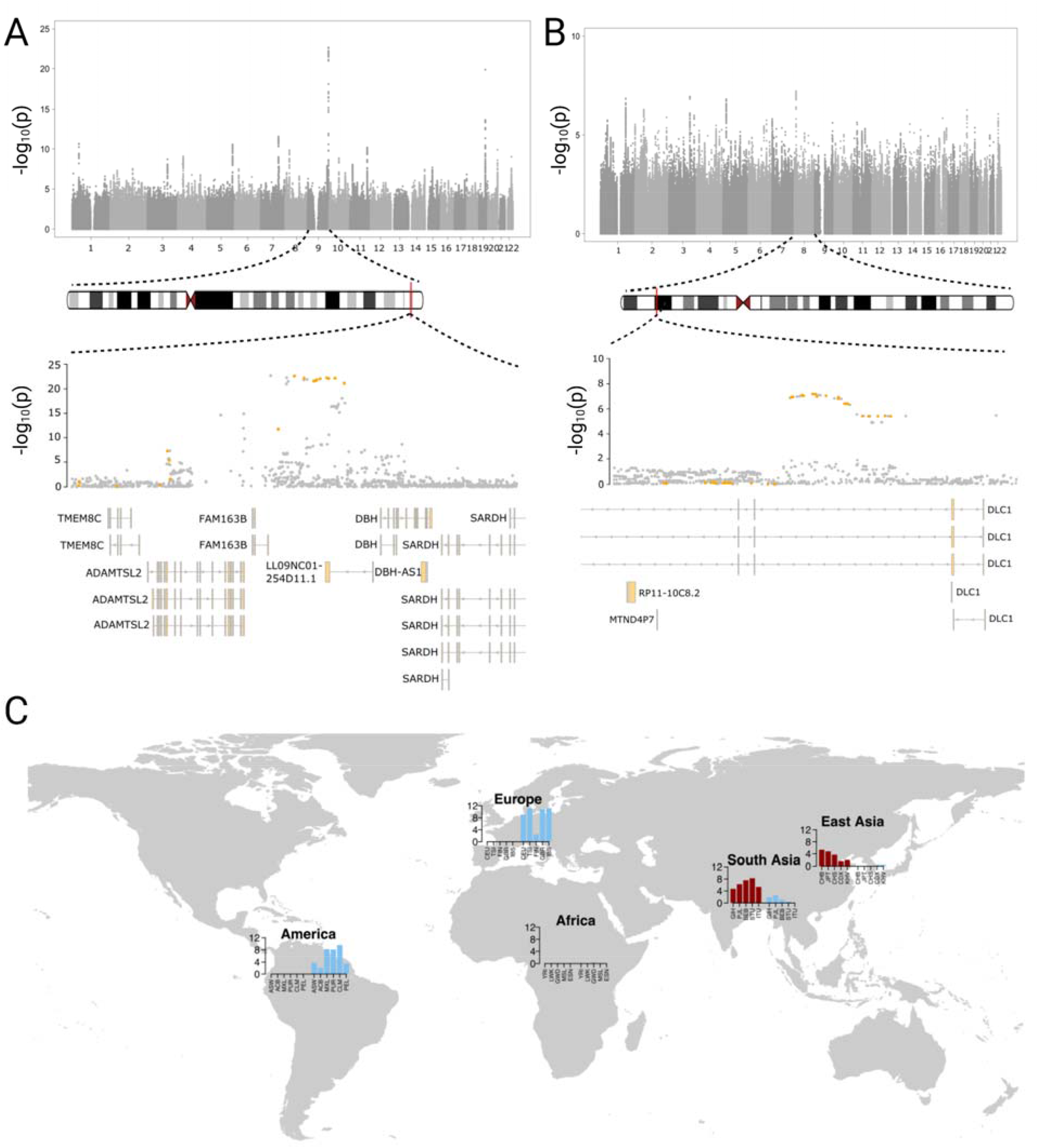
Top smoking risk loci in UK Biobank and Biobanks are linked to Neandertal DNA. **(A-B)** Manhattan plots for the ‘Current smoking status’ GWAS in the UK Biobank (panel A) and ‘Ever smoked’ GWAS in the Biobank Japan (panel B) are shown in the respective upper parts of the panels. A magnified view centered (+-100,000 base pairs) around the top genome-wide associations on chromosome 9 (A, chr9:136,363,019-136,563,019, P=2.7×10^−23^) and chromosome 8 (B, chr8:12,889,111-13,689,111, P=6.7×10^−8^) are displayed in the lower parts of panels A and B. Here, aSNPs are highlighted in orange. Overlapping genes and their gene models are shown in the lower part of the panel. **(C)** The frequency of the archaic alleles for the two top associated aSNPs (dark red: chr9:136,463,019; and light blue: chr8:13,289,111) in present-day 1,000 Genomes continental populations are displayed.

Importantly, 11 of the 18 Neandertal risk loci are linked to aSNPs with a frequency that was among the top 5% of aSNPs in at least one 1,000 Genomes population, including associations with all ten sleep-related traits, four of the five mental health phenotypes and two of the smoking habits. A particularly extreme example were aSNPs in the region of chr5:151,756,407-151,976,244, (association with chronotype and ‘getting up in the morning’) with frequencies between 21.5 and 55.2% in present-day Europeans, South Asians and Americans, putting them within the top 1% in 14 out of 15 of these populations. The aSNPs at this locus were also associated with modified expression of three genes (GLRA1, LINC01933 and NMUR2) in two brain regions and nerve tissue (Figure 4, Table S2). Archaic SNPs at another seven loci with links to four additional sleeping-related aSNP association, and one association for each, pain, smoking and mental health showed regulatory effects as well in various tissues including arteries, testis, thyroid, muscle spleen and ovaries (Table S2). In addition, we also found evidence for associated aSNPs affecting the amino acid sequences two genes; SCML4 (chr6:108,076,801_T/C, ‘period of unenthusiasm / disinterest’) and CHRNA5 (chr15:78885574_T/A, ‘Cigarette consumption per day’, Table S2).

**Figure 4:**
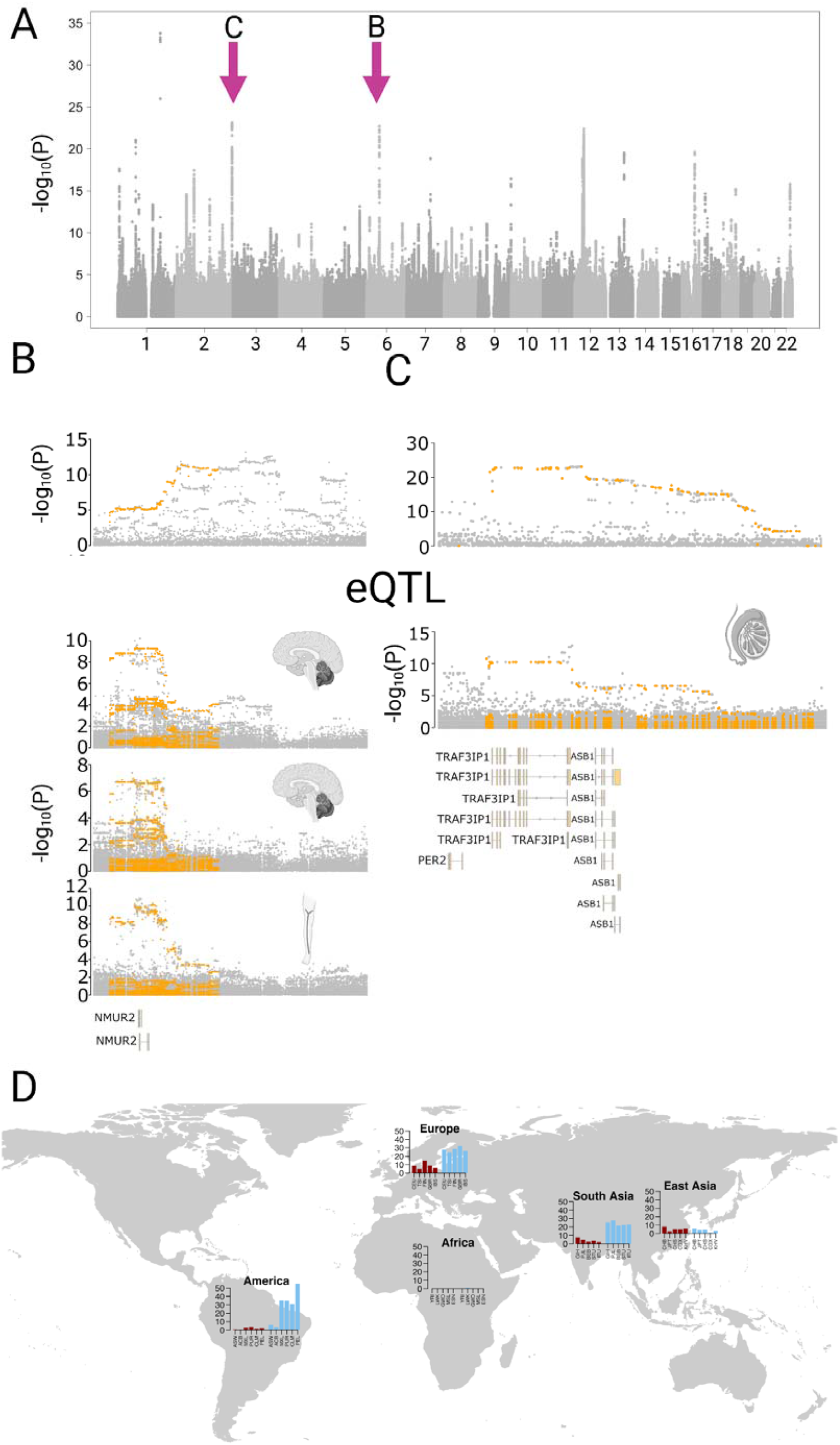
Neandertal DNA contributes to chronotype risk loci. **(A)** Manhattan plot for a Chronotype GWAS in the UK Biobank. **(B-C)** Magnified views of the association (y-axis, -log_10_ transformation of the association p value) score with chronotype on chromosomes 5 (B, chr5:151,589,813-152,689,813) and 2 (C, chr2:239,173,478-239,573,478) are shown on the top part of each panel. aSNPs are highlighted in orange. The middle part of each panel shows gene expression associations in GTEx tissues for the respective regions (eQTL -log^10^ transformed association P values for - from top - Cerebellar_Hemisphere, Cerebellum and Tibial Nerve in panel B and Testis in panel C). Models of overlapping gene are illustrated at the bottom of panels B and C. **(D)** The frequency of the top associated aSNPs from the two illustrated regions (chr2:239223478 in red and chr5:151889813 in blue) across 1,000 Genomes populations.

## Discussion

In this study we investigated the association strength of introgressed Neandertal DNA variants with neuropsychiatric disorders, nervous system medications (as a proxy for disease) and various behavioral (non-disease) phenotypes. Overall, while we found no associations with disease categories, there was an enrichment for associations of smoking and alcohol intake, pain and chronotype with Neandertal variants in the largest cohort (the UKBB). We also observed associations with some of these phenotypes in the NESDA and the Biobank Japan. Intriguingly, most of the enriched behavioral phenotypes closely resemble endophenotypes that are often strongly correlated with neuropsychiatric diseases, also on a genetic level. For example, recent work has suggested that there is a potentially genetic causal link between chronotype and odds of developing depressive symptoms(35) and similar findings have been reported for smoking(36) or alcohol abuse(37) and depression. In addition to smoking and alcohol intake, Neandertal DNA also showed significantly higher levels of association with pain and pain medications.

Our findings that introgressed variants are not enriched in psychiatric and neurological diagnostic categories are in line with a recent broad analysis that showed very limited associations of Neandertal variants with disease in modern humans beyond dermatological or immune-mediated disorders(38). The enrichment of associations with traits such as chronotype, pain, alcohol and tobacco use rather than diagnostic disease categories may thus reflect adaptations. Neandertal variants may thus persist in modern humans due to their neutral or even potentially advantageous effects at some point during recent evolution.

Interestingly, genomic regions that differ most between Neandertals and modern humans, including regions where no introgressed archaic DNA can be detected in people today(14,39) have previously been linked to brain-related genes (14,39). What exactly the evolutionary advantages of such phenotypes might have been, however, remains speculative at this point. Elevated frequencies of sleep-associated Neandertal variants are suggestive of them having been targets of adaptive processes. Sleep patterns and other behavioral phenotypes can be linked to circadian rhythm which in turn can be linked in differences of UV light exposure. If Neandertals were adapted to the UV light regime in Europe and Western Asia and contributed these adaptive alleles to modern humans, this may explain our enrichment results and in the case of sleep-related phenotypes suggest that they might have helped modern human adaptation to these new environments. With regard to pain, a previous analysis has indicated that Neandertal introgressed DNA near the SCN9A gene in modern humans is associated with lower pain thresholds and revealed a putative biological mechanism by implicating amino acid substitutions introduced by introgressed Neandertal variants in the sodium channel Nav1.7 protein(40). With regard to smoking or alcohol, evolutionary origins have been postulated for addiction, suggesting a co-evolution of the human brain, reward-seeking, and psychotropic substances(41). In line with this idea, “thrill seeking” was one of the phenotypes where we observed Neandertal associations in the UKBB. An alternative – non-exclusive - hypothesis could be that this reflects self-medication for pain(42), so that all of these associations (as well as the reported link with some pain medications) may be driven by the same selection processes.

Of note, in some instances, the enrichment results were driven by Neandertal variants that have reached genome-wide significance or may even - as in the case of smoking status in both the UK Biobank and Biobank Japan - represent the strongest association genome-wide. These may thus directly implicate certain biological pathways in the observed genetic associations. The majority of these genome-wide significant associations (18/27) were related to smoking and sleeping. Some of the association with sleep patterns were linked to Neandertal variants that have reached exceptionally high frequencies compared to other introgressed DNA in some present-day populations, suggesting that they may have been positively selected at some point in the past (Table S2). We also show that some of these variants are linked to well-established candidate genes for smoking status. For example, the aSNP chr6:108,076,801_T/C, which was associated with an increased ‘period of unenthusiasm / disinterest’ in our analysis of the UKBB, modifies an arginine to a glycine in the protein sequence of SCML4. This change is predicted as ‘probably damaging’ by PolyPhen(43) (Table S2). SCML4 has previously been linked to stress reactions in mice and modifications to its protein structure might therefore further contribute to related effects in mood phenotypes(44). Another protein sequence altering aSNP was chr15:78885574_T/A (archaic haplotype chr15:78,803,937-78,957,720), where the archaic allele A changes a histidine - the majority amino acid in present-day people - to a glutamine in CHRNA5 (Table S2). This modification is also classified as ‘possibly damaging’ by PolyPhen. CHRNA5 has been linked in several studies to smoking and various smoking risk factors(45–49).

In conclusion, our study provides an example of how evolutionary information can help interpreting the origin and genetic components of behavioral phenotypes. We show that while Neandertal DNA shows over-proportional numbers of associations with endophenotypes, this enrichment does not translate to disease. This evolutionary knowledge may help to decipher the environmental factors that shaped phenotypes.

## Data Availability

All data produced in the present study are available upon reasonable request to the authors

## Supplementary Tables

**Table S1: Neandertal DNA association enrichment results**

For each tested phenotype and for various significance cutoffs the enrichment results in the form of average ORs (with 95% CIs) and empirical P values are displayed.

**Table S2: Genome-wide significant Neandertal DNA risk loci**

Neandertal DNA associations with P<5×10^−8^ among the sets of tested phenotypes in this study, together with their associated GWAS summary statistics, inferred Neandertal haplotype, and overlapping eQTL and missense variants are provided.

**Table S3: Proportional Neandertal DNA association for groups of behavioral phenotypes**

The percentage of average odds ratio (OR) greater than one in eight groups of UK Biobank phenotype, the group of smoking phenotypes in the Biobank Japan and the group of tested NESDA phenotypes are provided. For each proportion of average ORs>1 a binomial CI together with its P value (tested against hypothesis of 50%) and false discovery rate (FDR) is available.

**Table S4: Classification of medication GWAS in the UK Biobank**

Medication GWAS from the UK Biobank are annotated based on three levels of WHO classifiers. This table contains only medication that was assigned to WHO nervous disease meta-classifier ‘N’.

## Acknowledgements

All figures were created with Biorender.com.

