## Supplementary Methods for "Neandertal introgression dissects the genetic landscape of neuropsychiatric disorders and associated behavioral phenotypes"

##### UK Biobank and its Neandertal DNA content

Summary statistics for genome-wide summary analysis (GWAS) from the UK Biobank(24) were obtained from the Neale lab [http://www.nealelab.is/uk-biobank/]. A detailed description of the analysis can be found at http://www.nealelab.is/blog/2017/9/11/details-and-considerations-of-the-uk-biobank-gwas and http://www.nealelab.is/blog/2019/10/24/updating-snp-heritability-results-from-4236-phenotypes-in-uk-biobank. In summary, GWAS have been conducted using 361,194 biobank individuals that have passed quality control filters. Based on these filters samples with non-British ancestry, are related, had sex chromosome aneuploidies or withdrawn from the biobank were excluded. Genotypes have been generated using two custom arrays and additional variants have been imputed using the 1,000 Genomes Project(29), the Haplotype Reference Consortium(50) and the UK10K cohort(51). The genotype data included ∼10.8 million SNPs with a minor allele frequency >0.1%, a Hardy–Weinberg equilibrium P value >1 × 10^−10^ and an INFO score, provided by the UK Biobank larger than 0.8. The association studies have been performed using the PHESANT software(52) and included as covariates the first 20 PCs, sex, age, age^2^, sex × age, and sex × age^2^ for diseases with male and female samples and only the nonsex covariates of this model for sex-specific diseases. We further restricted our analyses on 8,603,515 bi-allelic SNPs with a minor allele frequency larger than 1% in the cohort. Among those variants we identified 197,250 aSNPs.

##### Biobank Japan and its Neandertal DNA content

We used publicly available summary statistics for four smoking GWAS from the Biobank Japan(27). These GWAS have been conducted in ~200,000 individuals from the Biobank Japan cohort, with up to 165,436 individuals per GWAS. Included individuals have been between the ages of 20 and 89 and all been diagnosed with at least one of 45 diseases. Genotyping data has been generated using three custom arrays and SNPs characterized on multiple arrays included in the analysis. Samples with a high call rate (>0.98), unrelated and of East Asian ancestry have been kept. SNPs with a frequency below 0.005, call rate < 0.99 have been excluded. Imputation has been performed using 275 East Asians from the 1,000 Genomes and SNPs with a minor allele frequency below 1% and Hardy Weinberg equilibrium P < 1x10^-6^ were excluded. We included 5,826,586 of those SNPs into our analysis. We identified a total 62927 aSNPs among this set.

The smoking phenotypes have been defined as (1) ever versus never smokers, (2) smoking cessation, (3) age of smoking initiation and (4) quantity of smoking. The association analyses have been analyzed using a linear mixed model using phenotype-specific combinations of covariates including age, age^2^, sex and the status across 45 diseases.

##### NESDA GWAS generation and its Neandertal DNA content

Methods for biological sample collection and DNA extraction have been described previously(53) as well as quality control and imputation pipelines(54). Briefly, 95% of the samples were genotyped on the Affymetrix 6.0 Human SNP array and the remaining on the Perlegen-Affymetrix 5.0 array. After platform-specific QC the missing SNP genotypes between each platform were imputed using the GONL (Genome of the Netherlands)(55–57) reference panel and then merged, followed by additional more stringent QC.

This cross-platform GONL imputed dataset was used to identify ancestry outliers, defined based on Principal Components Analysis (PCA) by projecting 10 PCs from 1,000 Genomes reference set populations on the NTR cross-platform imputed data using the SMARTPCA program as described earlier(58,59). Individuals with PC values located outside of the range of European and/or British populations were defined as outliers. Upon exclusion of outliers, 10 PCs were recomputed for cross-platform imputed data to capture the variation within the Netherlands.

The SNPs from the cross-platform GONL imputed dataset (~1.3M) were used for a second round of imputations to the 1000G Phase 3(29) all ancestries reference panel using the Michigan Imputation Server(60).

Finally, the cross-platform imputed dataset was used to build a relationship matrix measuring genetic similarity using GCTA(61), which was pruned at 0.05 threshold in order to retain unrelated participants. Based on this genotype data we included 8,657,974 SNPs with a minor allele frequency larger than 1% for our analyses, 198,285 of which were aSNPs.

Genome-wide association analyses assuming an additive model were carried out using SNPTEST(28). All analyses were adjusted for age, sex and 10 ancestry-informative PCs. Association analyses related to alcohol intake were performed consistently with previous GWAS(62,63) including NESDA. The phenotype for this analyses was defined as the sex-specifc residuals (adjusted for age, age^2^, weight and 10 PCs) of (log_10_)gr/day alcohol intake.

### **Supplementary Methods References**
